# Rapid epidemic expansion of the SARS-CoV-2 Omicron variant in southern Africa

**DOI:** 10.1101/2021.12.19.21268028

**Authors:** Raquel Viana, Sikhulile Moyo, Daniel G Amoako, Houriiyah Tegally, Cathrine Scheepers, Christian L Althaus, Ugochukwu J Anyaneji, Phillip A Bester, Maciej F Boni, Mohammed Chand, Wonderful T Choga, Rachel Colquhoun, Michaela Davids, Koen Deforche, Deelan Doolabh, Susan Engelbrecht, Josie Everatt, Jennifer Giandhari, Marta Giovanetti, Diana Hardie, Verity Hill, Nei-Yuan Hsiao, Arash Iranzadeh, Arshad Ismail, Charity Joseph, Rageema Joseph, Legodile Koopile, Sergei L Kosakovsky Pond, Moritz UG Kraemer, Lesego Kuate-Lere, Oluwakemi Laguda-Akingba, Onalethatha Lesetedi-Mafoko, Richard J Lessells, Shahin Lockman, Alexander G Lucaci, Arisha Maharaj, Boitshoko Mahlangu, Tongai Maponga, Kamela Mahlakwane, Zinhle Makatini, Gert Marais, Dorcas Maruapula, Kereng Masupu, Mogomotsi Matshaba, Simnikiwe Mayaphi, Nokuzola Mbhele, Mpaphi B Mbulawa, Adriano Mendes, Koleka Mlisana, Anele Mnguni, Thabo Mohale, Monika Moir, Kgomotso Moruisi, Mosepele Mosepele, Gerald Motsatsi, Modisa S Motswaledi, Thongbotho Mphoyakgosi, Nokukhanya Msomi, Peter N Mwangi, Yeshnee Naidoo, Noxolo Ntuli, Martin Nyaga, Lucier Olubayo, Sureshnee Pillay, Botshelo Radibe, Yajna Ramphal, Upasana Ramphal, James E San, Lesley Scott, Roger Shapiro, Lavanya Singh, Pamela Smith-Lawrence, Wendy Stevens, Amy Strydom, Kathleen Subramoney, Naume Tebeila, Derek Tshiabuila, Joseph Tsui, Stephanie van Wyk, Steven Weaver, Constantinos K Wibmer, Eduan Wilkinson, Nicole Wolter, Alexander E Zarebski, Boitumelo Zuze, Dominique Goedhals, Wolfgang Preiser, Florette Treurnicht, Marietje Venter, Carolyn Williamson, Oliver G Pybus, Jinal Bhiman, Allison Glass, Darren P Martin, Andrew Rambaut, Simani Gaseitsiwe, Anne von Gottberg, Tulio de Oliveira

## Abstract

The severe acute respiratory syndrome coronavirus 2 (SARS-CoV-2) epidemic in southern Africa has been characterised by three distinct waves. The first was associated with a mix of SARS-CoV-2 lineages, whilst the second and third waves were driven by the Beta and Delta variants respectively^1–3^. In November 2021, genomic surveillance teams in South Africa and Botswana detected a new SARS-CoV-2 variant associated with a rapid resurgence of infections in Gauteng Province, South Africa. Within three days of the first genome being uploaded, it was designated a variant of concern (Omicron) by the World Health Organization and, within three weeks, had been identified in 87 countries. The Omicron variant is exceptional for carrying over 30 mutations in the spike glycoprotein, predicted to influence antibody neutralization and spike function^4^. Here, we describe the genomic profile and early transmission dynamics of Omicron, highlighting the rapid spread in regions with high levels of population immunity.

## Introduction

Since the onset of the COVID-19 pandemic in December 2019, variants of SARS-CoV-2 have emerged repeatedly. Some variants have spread worldwide and made major contributions to the cyclical infection waves that occur asynchronously in different regions. Between October and December 2020, the world witnessed the emergence of the first variants of concern (VOC). These variants exhibited increased transmissibility and/or immune evasion properties that threatened global efforts to control the pandemic. Although the Alpha, Beta and Gamma VOCs^2, 5^ that emerged during this time disseminated globally and drove epidemic resurgences in many different countries, it was the highly transmissible Delta variant that subsequently displaced all other VOC in most regions of the world^6^. During its spread, the Delta variant evolved into multiple sub-lineages^7^, some of which demonstrated signs of having a growth advantage in certain locations^8^, prompting speculation that the next VOC to drive a resurgence of infections would be likely derived from Delta. However, in October 2021, while Delta was continuing to exhibit high levels of transmission in the Northern hemisphere, a large Delta wave was subsiding in southern Africa. The culmination of this wave coincided with the emergence of a novel SARS-CoV-2 variant that, within days of its near-simultaneous discovery in four individuals in Botswana, a traveler from South Africa in Hong Kong, and 54 individuals in South Africa, was designated by the World Health Organization as Omicron: the fifth VOC of SARS-CoV-2.

## Results

### Epidemic dynamics and detection of Omicron

The three distinct epidemic waves of SARS-CoV-2 experienced by southern African countries were each driven by different variants: the first by descendants of the B.1 lineage^1^, the second by the Beta VOC^2, 9^, and the third by the Delta VOC^3^, with an estimated 2-5% of third wave cases in South Africa attributed to the C.1.2 lineage^10^ (Fig. 1A). Serosurveys conducted before the Delta wave suggested high levels of exposure to SARS-CoV-2 (40-60%) in South Africa^11, 12^, Malawi^13^, and Zimbabwe^14^, and modelled estimates suggested seroprevalence of 70-80% across South Africa by October 2021^15^. Accordingly, the weeks following the third wave in South Africa, between 10 October and 15 November 2021, were marked by a period of lower-level transmission as indicated by a low incidence of reported COVID-19 cases (100-200 new cases per day) and low (<2%) test positivity rates (Fig. 1A-1C).

**Figure 1:**
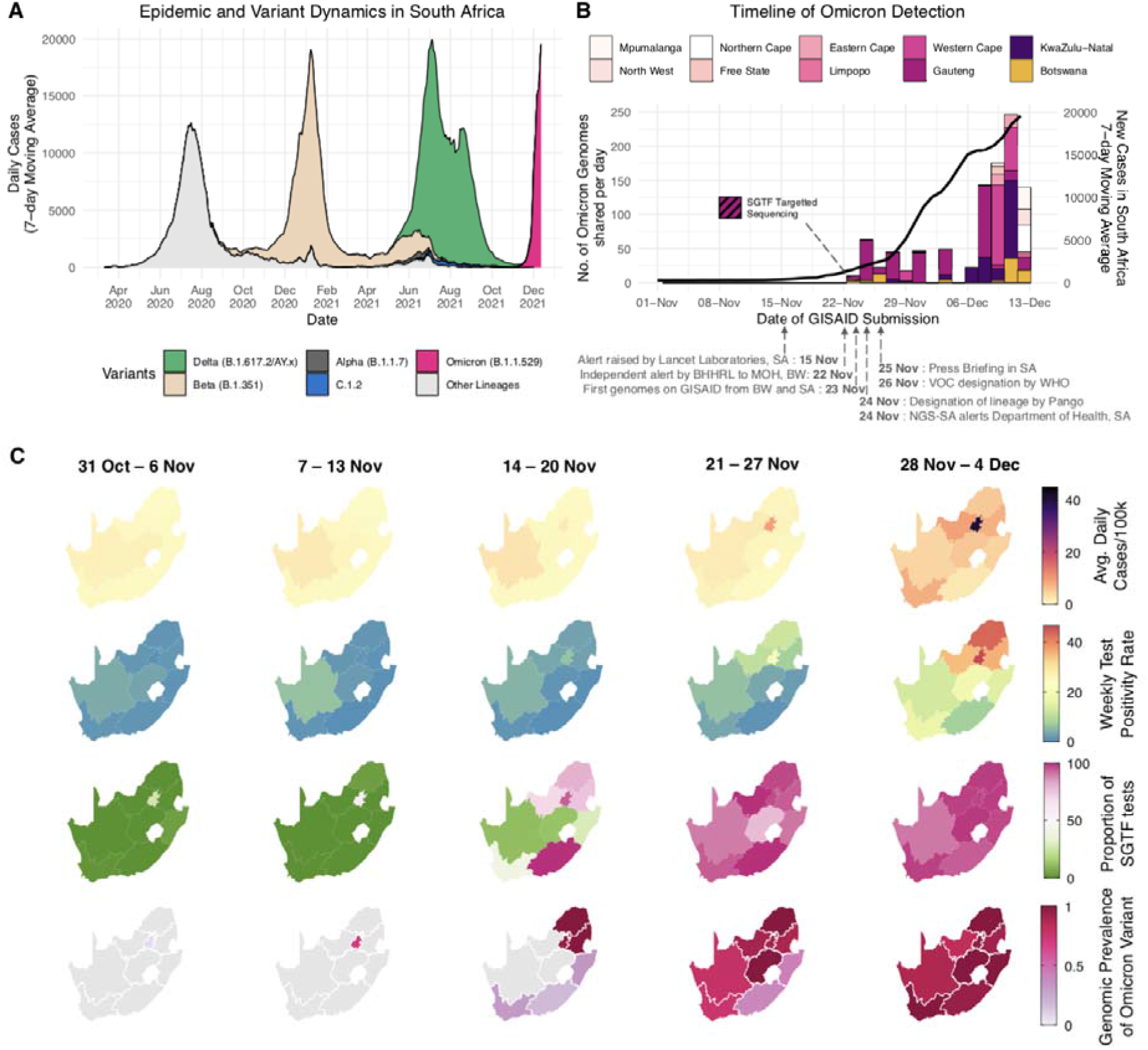
Detection of Omicron variant. A) The progression of daily reported cases in South Africa from March 2020 to December 2021. The 7-day rolling average of daily case numbers is coloured by the inferred proportion of variants responsible for the infections, as calculated by genomic surveillance data on GISAID. B) Timeline of Omicron detection in Botswana and South Africa. Bars represent the number of Omicron genomes shared per day, according to the date they were uploaded to GISAID, while the line represents the 7-day moving average of daily new cases in South Africa. C) Weekly progression of average daily cases per 100,000, test positivity rates, proportion of SGTF tests (on the TaqPath COVID-19 PCR assay) and genomic prevalence of Omicron in nine provinces of South Africa for five weeks from 31 October to 4 December 2021. Note that because of heterogeneous use of the TaqPath PCR assay across provinces, the proportion of SGTF tests illustrated for the Eastern Cape Province in weeks of 14 - 20 Nov and 21 - 27 Nov are based on only 2 and 4 data points respectively.

A rapid increase in COVID-19 cases was observed in mid-November 2021 in Gauteng province, the economic hub of South Africa containing the cities of Tshwane (Pretoria) and Johannesburg. Specifically, rising case numbers and test positivity rates were first noticed in Tshwane, initially associated with outbreaks in higher education settings. This resurgence of cases was accompanied by an increasing frequency of S-gene target failure (SGTF) during TaqPath-based (Thermo Fisher Scientific) diagnostic PCR testing: a phenomenon previously observed with the Alpha variant due to a deletion at amino acid positions 69 and 70 (Δ69-70) in the SARS-CoV-2 spike protein. Given the low prevalence of Alpha in South Africa (Fig. 1A), targeted whole-genome sequencing of these specimens was prioritized.

On 19 November 2021, sequencing results of an initial batch of 8 SGTF samples collected between 14-16 November 2021 indicated that all were a new and genetically-distinct lineage of SARS-CoV-2. Further rapid sequencing identified the same variant in 29 of 32 routine diagnostic samples from multiple locations in Gauteng Province, indicating widespread circulation of this new variant by the second week of November. Crucially, this rise immediately preceded a sharp increase in reported case numbers (Fig. 1C, Extended Data Fig. 1). In the following four days this lineage was confirmed by sequencing in another two provinces: KwaZulu-Natal (KZN) and the Western Cape (Fig. 1B).

Concurrently in Gaborone, Botswana (∼360km from Tshwane), four genomes generated from samples collected on 11 November 2021, and sequenced on 17-18 November 2021 as part of weekly surveillance, displayed an unusual set of mutations. These were reported to the Botswana Ministry of Health and Wellness on 22 November 2021, as “unusual sequences” that were linked to a group of visitors (non-residents) on a diplomatic mission. The sequences were uploaded to GISAID^17, 18^ on 23 November 2021, and it became apparent that they belonged to a new lineage. A further 15 genomically confirmed cases (not epidemiologically linked to the first four) were identified within the same week from various other locations in Botswana. All of these either had travel links from South Africa, or were contacts of someone with travel links.

On 24 November 2021, these SARS-CoV-2 genomes from both South Africa and Botswana were designated as belonging to a new PANGO lineage (B.1.1.529)^19^, later divided into sub-lineages aliased BA.1 (the main clade), BA.2 and BA.3. On 26 November 2021, the lineage was designated a VOC and named Omicron by the WHO on the recommendation of the Technical Advisory Group on SARS-CoV-2 Virus Evolution^20^. By the first week of December 2021, Omicron was causing a rapid and sustained increase in cases in South Africa and Botswana (Fig. 1C, Extended Data Fig. 2 for Botswana). In Gauteng, weekly test positivity rates increased from <1% in the week beginning 31 October, to 16% in the week beginning 21 November 2021, and to 35% in the week beginning 28 November, concurrently with an exponential rise in COVID-19 incidence (Fig. 1C, Extended Data Fig. 1). Nationally, daily case numbers exceeded 22 000 (84% of the peak of the previous wave of infections) by 9 December 2021. At the same time, the proportion of TaqPath PCR tests with SGTF increased rapidly in all provinces of South Africa reaching ∼90% nationally by the week beginning 21 November 2021, strongly indicating that the fourth wave was being driven by Omicron: an indication that has now been confirmed by virus genome sequencing in all provinces (Fig. 1C). Similarly, Botswana experienced a sharp increase in cases, doubling every 2-3 days late November to early December 2021, transitioning from a 7-day moving average of <10 cases/100 000 to above 25 cases/100,000 in less than 10 days (Extended Data Fig. 2).

By 16 December 2021, Omicron had been detected in 87 countries, both in samples from travelers returning from southern Africa, and in samples from routine community testing (Extended Data Fig. 3).

### The evolutionary origins of Omicron

To determine when and where Omicron likely originated, we analyzed all 686 available Omicron genomes (including 248 from southern Africa and 438 from elsewhere in the world) retrieved from GISAID (date of access 7 December 2021)^17, 18^, in the context of a global reference set of representative SARS-CoV-2 genomes (n=12 609) collected between December 2019 and November 2021. Preliminary maximum-likelihood phylogenies identified the BA.1/Omicron sequences as a monophyletic clade rooted within the B.1.1 lineage (Nextstrain clade 20B), with no clear basal progenitor (Fig. 2A). Importantly, the BA.1/Omicron cluster is highly phylogenetically distinct from any known VOC or variants of interest (VOI) and from any other lineages known to be circulating in southern Africa (e.g. C.1.2) (Fig. 2A). More recently, two related lineages have emerged (BA.2 and BA.3), both sharing many, but not all of the characteristic mutations of BA.1/Omicron and both having many unique mutations of their own. We primarily focus here on the BA.1 lineage which is rapidly spreading in multiple countries around the world and is the lineage first officially designated as the Omicron VOC.

**Figure 2:**
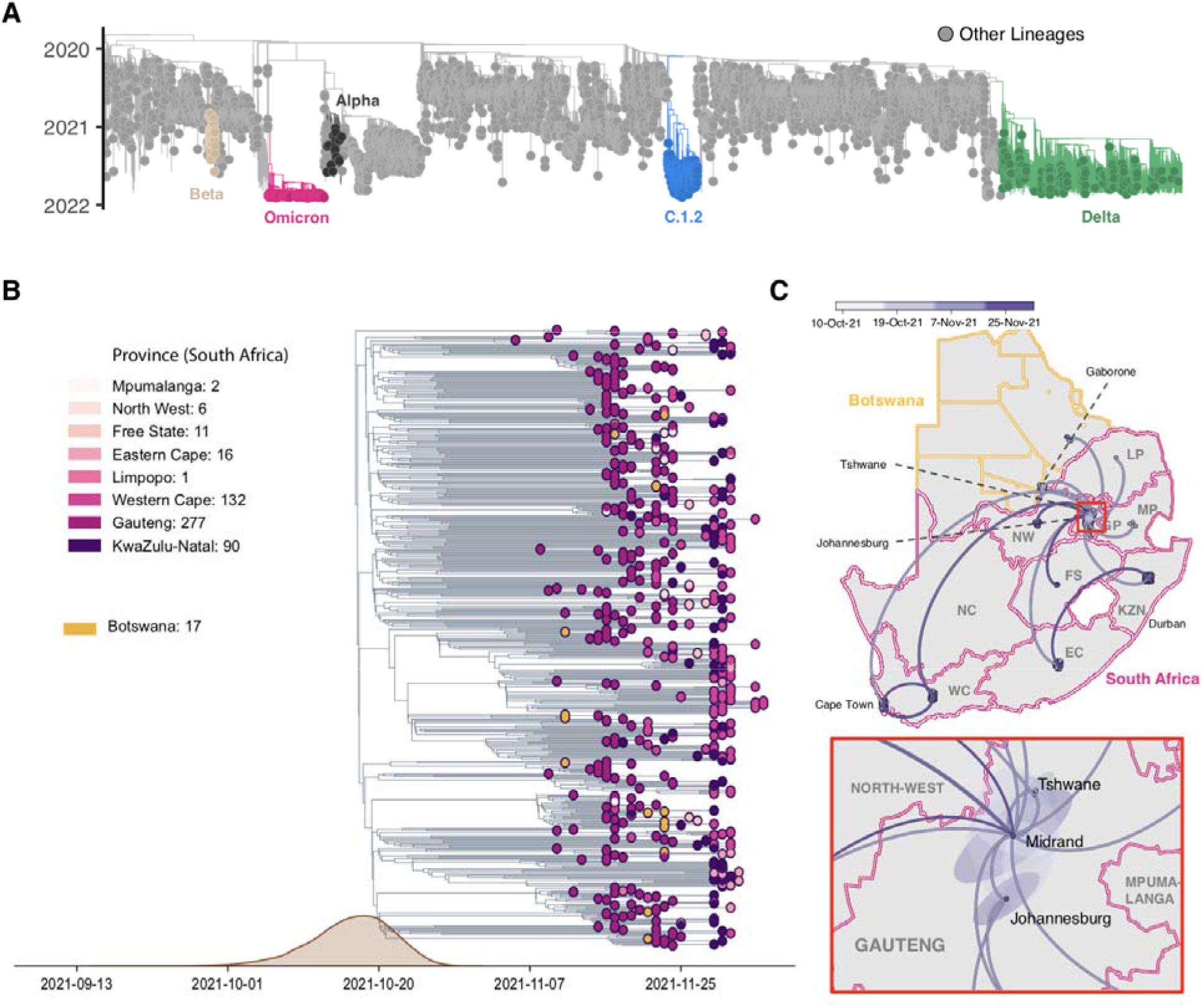
Evolution of Omicron. A) Time-resolved maximum likelihood phylogeny of 13,295 SARS-CoV-2 sequences; 9,944 of these are from Africa (denoted with tip point circle shapes). Alpha, Beta and Delta VOCs and the C.1.2 lineage, recently circulating in South Africa, are denoted in black, brown, green and blue respectively. The newly identified SARS-CoV-2 Omicron variant is shown in pink. Genomes of other lineages are shown in grey. B) Time-resolved maximum clade credibility phylogeny of the Omicron cluster of southern African genomes (n = 553), with locations indicated. The distribution of estimated time of origin is also shown. C) Spatiotemporal reconstruction of the spread of the Omicron variant in Southern Africa with an inset of Gauteng province. Circles represent nodes of the maximum clade credibility phylogeny, coloured according to their inferred time of occurrence (scale in top panel). Shaded areas represent the 80% highest posterior density interval and depict the uncertainty of the phylogeographic estimates for each node. Solid curved lines denote the links between nodes and the directionality of movement is anticlockwise along the curve.

Time-calibrated Bayesian phylogenetic analysis of all BA.1 assigned genomes from southern Africa (as of 11 December 2021, n=553) estimated the time when the most recent common ancestor of the analysed BA.1 lineage sequences existed to be 9 October 2021 (95% credible intervals 30 September - 20 October) with a per-day exponential growth rate of 0.136 (95% confidence interval (CI) 0.100 – 0.173) reflecting a doubling time of 5.1 days (95% CI 4.0 – 6.9) (Fig 2B). These estimates are robust to whether the evolutionary rate is estimated from the data or fixed to previously estimated values (Extended Data Table S1). Limiting the analysis to a subset of genomes from Gauteng Province only (279 genomes) yields a faster growth rate estimate with a doubling time of 1.8 days (95% CI 1.4 – 3.0) (**Extended Data S1**). Using a phylodynamic model that accounts for variable genome sampling through time (birth-death skyline model) yields doubling times of Omicron in South Africa between 3.88 and 3.99 days and mean effective reproduction number estimates (Re) of 2.74 to 2.79 for the period of early November to early December (Extended Data Table X). Spatiotemporal phylogeographic analysis indicates that the BA.1/Omicron variant spread from the Gauteng province of South Africa to seven of the eight other provinces and to two regions of Botswana from late October to late November 2021, and shows evidence of more recent transmission within and between other South African provinces (Fig 2C).

### Molecular profile of Omicron

Compared to Wuhan-Hu-1, Omicron carries 15 mutations in the spike receptor-binding domain (RBD) (Fig. 3), five of which (G339D, N440K, S477N, T478K, N501Y) have been shown individually to enhance hACE2 binding^21^. Seven of the RBD mutations (K417N, G446S, E484A, Q493R, G496S, Q498R and N501Y) are expected to have moderate to strong impacts on binding of at least three of the four major classes of RBD-targeted neutralizing antibodies (NAbs)^22–24^. These RBD mutations coupled with four amino acid substitutions (A67V, T95I, G142D, and L212I), three deletions (69-70, 143-145 and 211) and an insertion (EPE between 214 and 215) in the N-terminal domain (NTD)^25^, are predicted to underlie the substantially reduced sensitivity of Omicron to neutralization by anti-SARS-CoV-2 antibodies induced by either infection or vaccination^26, 27^. These mutations also involve key structural epitopes targeted by some of the currently authorized monoclonal antibodies, particularly bamlanivimab + etesevimab and casirivimab + imdevimab^28–30^. Preliminary analysis suggests that although the spike mutations involve a number of T cell and B cell epitopes, the majority of epitopes (>70%) remain unaffected^31^.

**Figure 3.**
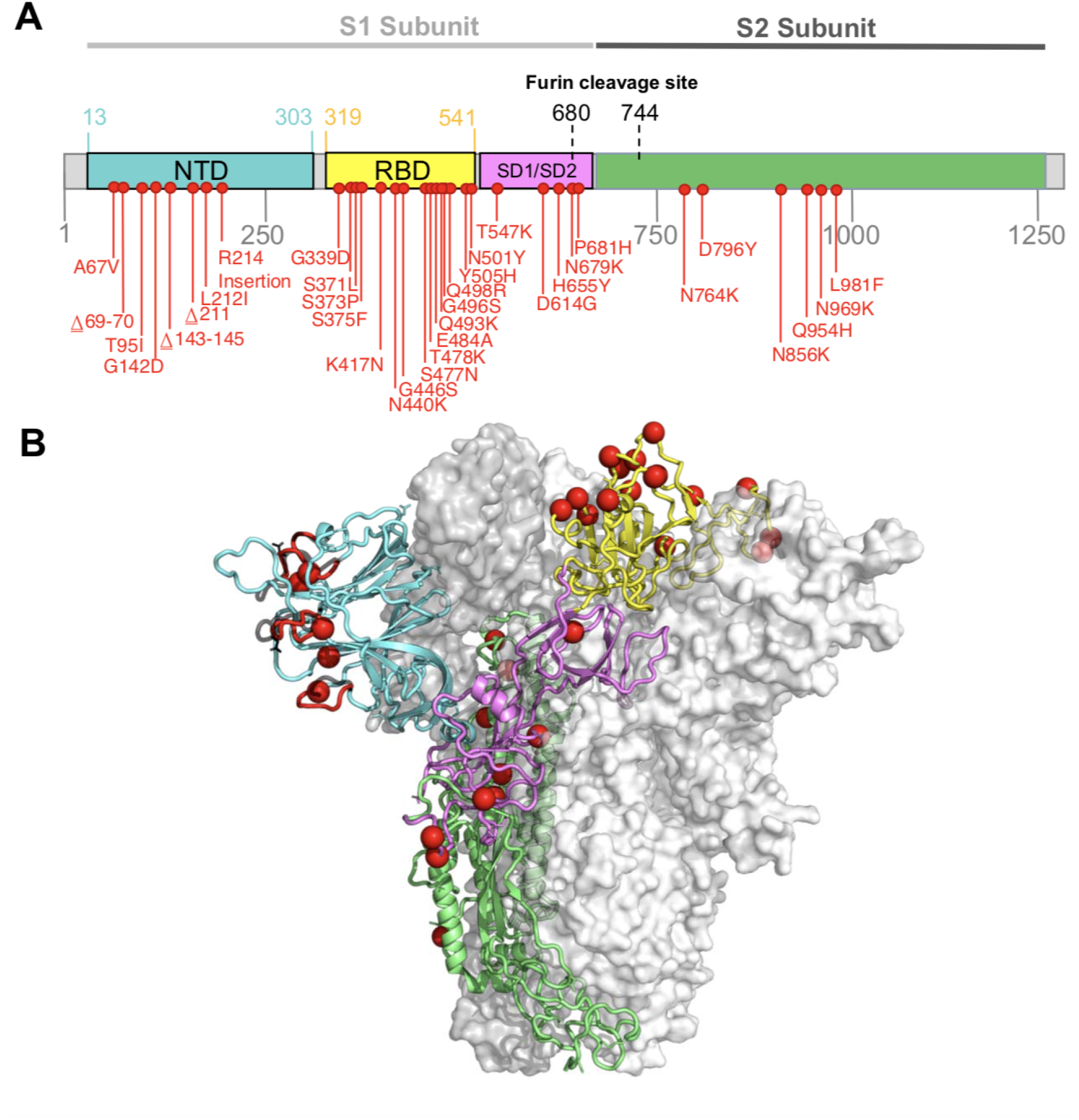
Molecular profile of Omicron. A) Amino-acid mutations on the spike gene of the BA.1/Omicron variant. B) Structure of the SARS-CoV-2 Spike trimer, showing a single spike protomer in cartoon view. The N terminal domain, receptor binding domain, subdomains 1 and 2, and the S2 protein are shown in cyan, yellow, pink, and green respectively. Red spheres indicate the alpha carbon positions for each omicron variant residue. NTD-specific loop insertions/deletions are shown in red, with the original loop shown in transparent black.

Omicron also has a cluster of three mutations (H655Y, N679K and P681H) adjacent to the S1/S2 furin cleavage site (FCS) which are likely to enhance spike protein cleavage and fusion with host cells^32, 33^ and which could also contribute to enhanced transmissibility^34^ (Extended Data Fig. 4).

Outside of the spike protein, a deletion in nsp6 (105-107del), in the same region as deletions seen in Alpha, Beta, Gamma and Lambda, may have a role in evasion of innate immunity^35^; and the double mutation in nucleocapsid (R203K, G204R), also present in Alpha, Gamma and C.1.2, has been associated with enhanced infectivity in human lung cells ^36^.

### Omicron is not obviously recombinant

Given the large number of mutations differentiating BA.1/Omicron and BA.2 from other known SARS-CoV-2 lineages it was considered plausible that either (i) both of these lineages might have descended from a common recombinant ancestor, or (ii) that one of the BA lineages might have originated via recombination between a virus in the other BA lineage and a virus in a non-BA lineage. We tested these hypotheses using a variety of recombination detection approaches (implemented in the programs GARD^37^; 3SEQ^38^; and RDP5^39^) to identify potential signals of recombination in sequence datasets containing BA.1 and BA.2 sequences together with sequences representative of global SARS-CoV-2 genomic diversity.

Potential evidence of recombination was identified by 3SEQ, GARD and RDP5 in these datasets. The most likely recombination breakpoint locations were located between nucleotide positions 20520 and 21619 (near the start of the S-gene; supported by GARD, RDP5 and 3SEQ) and between 23609 and 23614 (near the middle of the S-gene; supported by GARD and 3SEQ). 3SEQ identified an additional breakpoint at nucleotide position 24513 (toward the end of the S-gene). Phylogenetic analysis of the genome regions bounded by these breakpoints (genome coordinates 1450-20520, 21619-23609 and 23614-24513) revealed no support for a recombinant origin for either the BA.1 or BA.2 lineages (Extended Data Fig. 5). Although one BA.1 isolate (Botswana/R43B66) displayed evidence of having potentially inherited nucleotides 23614-24513 from a Delta virus by recombination, there was no strong phylogenetic support for the clustering of this sequence with Delta viruses. Further, read coverage in this region of the Botswana/R43B66 sequence was so low that we were unable to exclude the possibility that the apparent recombination signal was attributable to a combination of miscalled/uncalled nucleotides and alignment uncertainty.

Although we found no convincing phylogenetic or statistical evidence of either the most recent common ancestor of BA.1 and BA.2 being recombinant, or of the most recent common ancestors of either the BA.1 or BA.2 lineages having been derived through recombination, it should be noted that recombination tests in general will not have sufficient statistical power to reliably identify evidence of individual recombination events that result in transfers of less than ∼5 contiguous polymorphic nucleotide sites between genomes^37, 40, 41^. Further, if BA.1 and/or BA.2 are the products of a series of multiple partially overlapping recombination events occurring across multiple temporally clustered replication cycles, the complex patterns of nucleotide variation that might result could be extremely difficult to interpret as recombination using the methods applied here^42^.

### Signs of strong selection and epistasis during the origin and ongoing evolution of the Omicron lineage

We applied a selection analysis pipeline to all available sequences designated as BA.1 in GISAID as of 8 December 2021. The analysis followed the procedure described previously^35^, and downsampled alignments of individual protein encoding regions to obtain a median of 25 unique Omicron haplotype sequences and 107 unique haplotype sequences for each gene/ORF from a representative selection of other SARS-CoV-2 lineages (used as background sequences to contextualize evolution within the Omicron sub-clade).

We detected evidence of gene-wide positive selection (using the BUSTED method^43^) acting on six genes/ORFs since the ancestral BA.1/Omicron and BA.2 lineage split from the B.1.1 lineage: S-gene (p < 0.0001), exonuclease (p < 0.0001), nsp6 (p = 0.001), M-gene (p = 0.002), N-gene (p = 0.006), and E-gene (p = 0.05) In all six genes, this selection was strong (dN/dS > 10), and occurred in bursts (≤ 6% of branch/site combinations selected). The branch separating BA.1/Omicron from its most recent B.1.1 ancestor had the most prominent selection signal (which was strongest in the S-gene; BUSTED p-value < 0.0001 with dN/dS > 100 at ∼0.5% of S-gene codon sites^44^), strongly supporting the hypothesis that adaptive evolution played a significant role in the mutational divergence of Omicron from other B.1.1 SARS-CoV-2 lineages. Relative to the intensity of selection evident within the background B.1.1 lineages, selection in three genes was likely significantly intensified in the ancestral Omicron lineage: S-gene (intensification factor K = 2.0; p < 0.0001 ^45^), exonuclease (K = 4.0; p < 0.0001), and nsp6 (K = 5.1; p = 0.02).

Among 294 codon sites that are polymorphic among the BA.1/Omicron sequences analysed, 32 were found to have experienced episodic positive selection since BA.1 split from the B.1.1 lineage (MEME p ≤ 0.01, Extended Data Table S2). Sixteen (50%) of these codon sites are in the S-gene, 13 of which contain BA.1 lineage-defining mutations (i.e. these selection signals reflect mutations that occurred within the ancestral Omicron lineage). The three positively selected codon sites that did not correspond to sites of lineage-defining mutations (S/346, S/452, and S/701) are particularly notable as these are attributable to mutations that have occurred since the MRCA of the analysed BA.1 sequences. The mutations driving the positive selection signals at these three sites in the Omicron S-gene converge on mutations seen in other VOCs or VOIs (R346K in Mu, L452R in Delta, and A701V in Beta and Iota). The A701V mutation, the precise impact of which is currently unknown, is one of 19 in a proposed “501Y lineage Spike meta-signature” comprising the set of mutations that were most adaptive during the evolution of the Alpha, Beta and Gamma VOC lineages^35^. Further, both R346K and L452R are known to impact antibody binding^23^ and both of the codon sites where these mutations occur display evidence for directional selection (using the FADE method^47^). These selective patterns suggest that, during its current explosive spread, Omicron may be undergoing additional evolution to modify its neutralization profile.

### Potential for increased transmissibility and immune evasion

We estimated that Omicron had a growth advantage of 0.24 (95% CI: 0.16-0.33) per day over Delta in Gauteng, South Africa (Fig. 4A). This corresponds to a 5.4-fold (95% CI: 3.1-10.1) weekly increase in cases compared to Delta. The growth advantage of Omicron is likely to be mediated by (i) an increase relative to other variants of its intrinsic transmissibility, (ii) an increase relative to other variants in its capacity to infect, and be transmitted from, previously infected and vaccinated individuals; or (iii) both.

**Figure 4:**
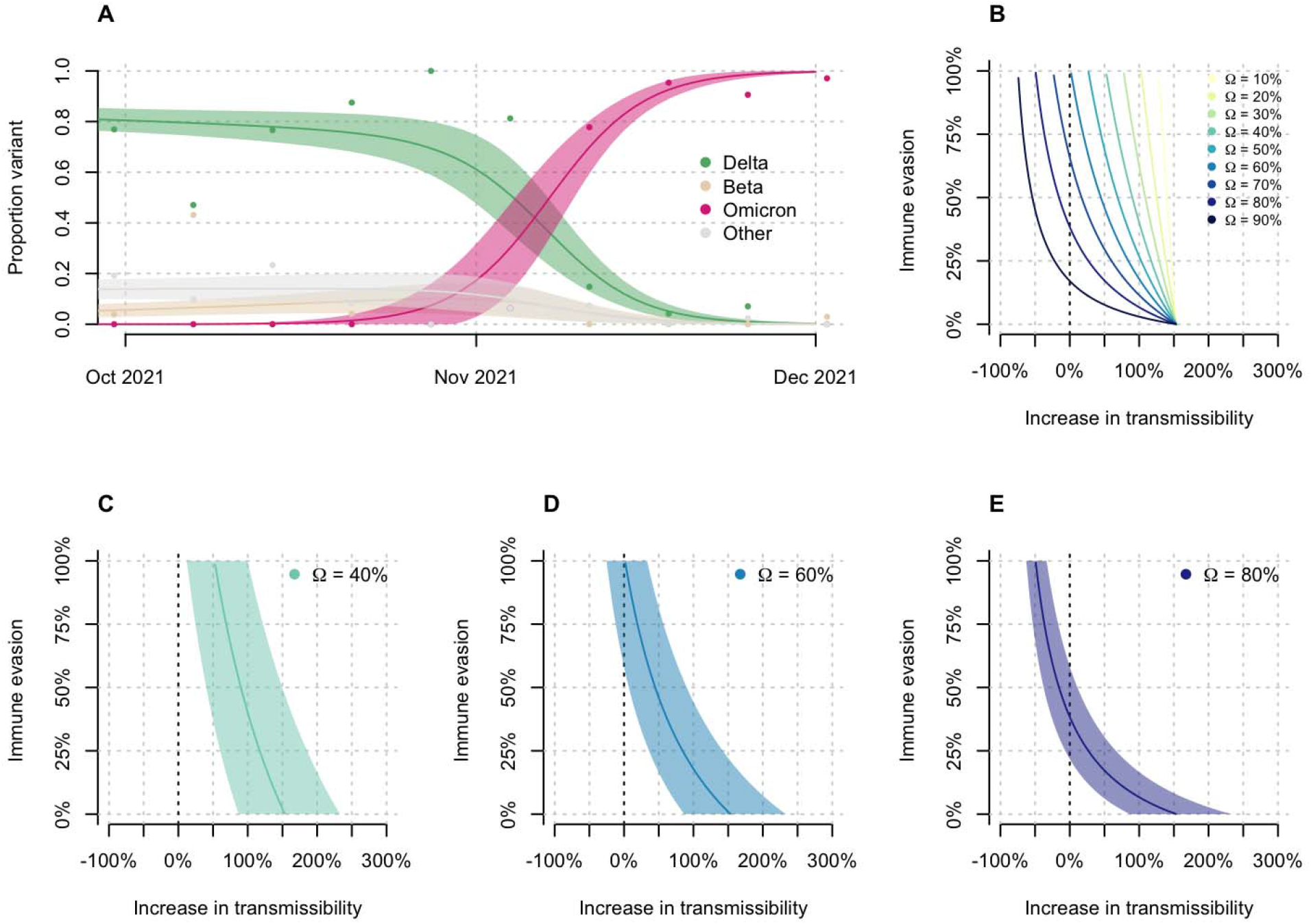
Growth of Omicron in Gauteng, South Africa, and relationship between potential increase in transmissibility and immune evasion. (A) Omicron rapidly outcompeted Delta in November 2021. Model fits are based on a multinomial logistic regression. Dots represent the weekly proportions of variants. (B) The relationship between the potential increase in transmissibility and immune evasion strongly depends on the assumed level of current population immunity against Delta (Ω). (C-E) Relationship for a population immunity of 40%, 60%, and 80% against infection and transmission with Delta. The dark vertical dashed line indicates equal transmissibility of Omicron compared to Delta. Shaded areas correspond to the 95% CIs of the model estimates.

The predicted combination of transmissibility and immune evasion for Omicron strongly depends on the assumed level of current population immunity against infection by, and transmission of, the competing variant Delta that is afforded by prior-infections with wild-type Wuhan, Beta, Delta, and other strains, and/or vaccination (Fig. 4B). For moderate levels of population immunity against Delta (Ω = 0.4), immune evasion alone cannot explain the observed growth advantage of Omicron (Fig. 4C). For medium levels of immunity against Delta (Ω = 0.6), very high levels of immune evasion could explain the observed growth advantage without an additional increase in transmissibility (Fig. 4D). For high levels of population immunity against Delta (Ω = 0.8), even moderate levels of immune evasion (∼25-50%) can explain the observed growth advantage without an additional increase in transmissibility (Fig. 4E). The results of seroprevalence studies and vaccination coverage (∼40% of the adult population) in South Africa suggest that the proportion of the population with potential immunity against Delta and earlier variants is likely to be above 60%. We thus argue that the population level of protective immunity against Delta is high, and that partial immune evasion is a major driver for the observed dynamics of Omicron in South Africa. This notion is supported by recent findings that show an increased risk of SARS-CoV-2 reinfection associated with the emergence of Omicron in South Africa^48^ and the initial results from neutralization assays^26, 27^. In addition to immune evasion, an increase, or decrease, in the transmissibility of Omicron compared to Delta cannot, however, be ruled out.

There are a number of limitations to this analysis. First, we estimated the growth advantage of Omicron based on early sequence data only. These data could be biased due to targeted sequencing of SGTF samples and stochastic effects (e.g., superspreading) in a low incidence setting, which can lead to overestimates of the growth advantage, and consequently of the increased transmissibility and immune evasion. Second, without reliable estimates of the level of protective immunity against Delta in South Africa, we cannot obtain precise estimates of transmissibility or immune evasion of Omicron.

## Conclusion

Strong genomic surveillance systems in South Africa and Botswana enabled the identification of Omicron within a week of observing a resurgence in cases in Gauteng Province. Immediate notification of the WHO and early designation as a VOC has stimulated global scientific efforts and has given other countries time to prepare their response. Omicron is now driving a fourth wave of the SARS-CoV-2 epidemic in southern Africa, and is spreading rapidly in several other countries. Genotypic and phenotypic data suggest that Omicron has the capacity for substantial evasion of neutralizing antibody responses, and modelling suggests that immune evasion could be a major driver of the observed transmission dynamics. Close monitoring of the spread of Omicron in countries outside southern Africa will be necessary to better understand its transmissibility and the capacity of this variant to evade post-infection and vaccine-elicited immunity. Neutralizing antibodies are only one component of the immune protection from vaccines and prior infection, and the cellular immune response is predicted to be less affected by the mutations in Omicron. Vaccination therefore remains critical to protect those at highest risk of severe disease and death. The emergence and rapid spread of Omicron poses a threat to the world and a particular threat in Africa, where fewer than one in ten people is fully vaccinated.

## Data Availability

All genomes generated in this research at public available at GISAID. The short reads are public available at the Short Read Archives of NCBI.

## Methods

### Epidemiological dynamics

We analyzed daily cases of SARS-CoV-2 in South Africa up to 14 December 2021 from publicly released data provided by the National Department of Health and the National Institute for Communicable Diseases. This was accessible through the repository of the Data Science for Social Impact Research Group at the University of Pretoria (https://github.com/dsfsi/covid19za)^49,50^. The National Department of Health releases daily updates on the number of confirmed new cases, deaths and recoveries, with a breakdown by province. Daily case numbers for Botswana were obtained via Our World in Data (OWID) COVID-19 data repository (https://github.com/owid/covid-19-data). We consulted estimates for the *R*e of SARS-CoV-2 in South Africa and Botswana from the ‘covid-19-Re’ data repository (https://github.com/covid-19-Re/dailyRe-Data)^51^. We obtained test positivity data from weekly reports from the National Institute for Communicable Diseases (NICD)^52^. Data to calculate the proportion of positive Thermo Fisher TaqPath COVID-19 PCR tests with SGTF in South Africa was obtained from the National Health Laboratory Service and Lancet Laboratories. Test positivity data for Botswana was obtained from the National Health Laboratory through 6 December 2021. All data visualization was generated through the ggplot package in R^53^.

### SARS-CoV-2 sampling

As part of the NGS-SA, seven sequencing hubs in South Africa receive randomly selected samples for sequencing every week according to approved protocols at each site^54^. These samples include remnant nucleic acid extracts or remnant nasopharyngeal and oropharyngeal swab samples from routine diagnostic SARS-CoV-2 PCR testing from public and private laboratories in South Africa. In response to a focal resurgence of COVID-19 in the City of Tshwane Metropolitan Municipality in Gauteng Province in November, we enriched our routine sampling with additional samples from the affected area, including initial targeted sequencing of SGTF samples. In Botswana, all public and private laboratories submit randomly selected residual nasopharyngeal and oropharyngeal PCR positive samples weekly to the National Health Laboratory (NHL) and the Botswana Harvard HIV Reference Laboratory (BHHRL) for sequencing.

### Ethical statement

The genomic surveillance in South Africa was approved by the University of KwaZulu–Natal Biomedical Research Ethics Committee (BREC/00001510/2020), the University of the Witwatersrand Human Research Ethics Committee (HREC) (M180832), Stellenbosch University HREC (N20/04/008_COVID-19), University of Cape Town HREC (383/2020), University of Pretoria HREC (H101/17) and the University of the Free State Health Sciences Research Ethics Committee (UFS-HSD2020/1860/2710). The genomic sequencing in Botswana was conducted as part of the national vaccine roll-out plan and was approved by the Health Research and Development Committee (Health Research Ethics body, HRDC#00948 and HRDC#00904). Individual participant consent was not required for the genomic surveillance. This requirement was waived by the Research Ethics Committees.

### Ion Torrent Genexus Integrated Sequencer methodology for rapid whole genome sequencing of SARS-CoV-2

Viral RNA was extracted using the MagNA Pure 96 DNA and Viral Nucleic Acid kit on the automated MagNA Pure 96 system (Roche Diagnostics, USA) as per the manufacturer’s instructions. Extracts were then screened by qPCR to acquire the mean cycle threshold (Ct) values for the SARS-CoV-2 N-gene and ORF1ab-gene using the TaqMan 2019-nCoV assay kit v1 (ThermoFisher Scientific, USA) on the ViiA7 Real-time PCR system (ThermoFisher Scientific, USA) as per the manufacturer’s instructions. Extracts were sorted into batches of N=8 within a Ct range difference of 5 for a maximum of two batches per run. Extracts with <200 copies were sequenced using the low viral titer protocol. Next-generation sequencing was performed using the Ion AmpliSeq SARS-CoV-2 Research Panel on the Ion Torrent Genexus Integrated Sequencer (ThermoFisher Scientific, USA) which combines automated cDNA synthesis, library preparation, templating preparation and sequencing within 24 hours. The Ion Ampliseq SARS-CoV-2 Research Panel consists of 2 primer pools targeting 237 amplicons tiled across the SARS-CoV-2 genome providing >99% coverage of the SARS-CoV-2 genome (∼30 kb) and an additional 5 primer pairs targeting human expression controls. The SARS-CoV-2 amplicons range from 125 to 275 bp in length. TRINITY was utilised for de novo assembly and the Iterative Refinement Meta-Assembler (IRMA) for genome assisted assembly as well as FastQC for quality checks.

### Whole-genome sequencing and genome assembly

RNA was extracted on an automated Chemagic 360 instrument, using the CMG-1049 kit (Perkin Elmer, Hamburg, Germany). The RNA was stored at −80 ◦C prior to use. Libraries for whole genome sequencing were prepared using either the Oxford Nanopore Midnight protocol with Rapid Barcoding or the Illumina COVIDseq Assay.

### Illumina Miseq/NextSeq

For the Illumina COVIDseq assay, the libraries were prepared according to the manufacturer’s protocol. Briefly, amplicons were tagmented, followed by indexing using the Nextera UD Indexes Set A. Sequencing libraries were pooled, normalized to 4 nM and denatured with 0.2 N sodium acetate. A 8 pM sample library was spiked with 1% PhiX (PhiX Control v3 adaptor-ligated library used as a control). We sequenced libraries on a 500-cycle v2 MiSeq Reagent Kit on the Illumina MiSeq instrument (Illumina). On the Illumina NextSeq 550 instrument, sequencing was performed using the Illumina COVIDSeq protocol (Illumina Inc, USA), an amplicon-based next-generation sequencing approach. The first strand synthesis was carried using random hexamers primers from Illumina and the synthesized cDNA underwent two separate multiplex PCR reactions. The pooled PCR amplified products were processed for tagmentation and adapter ligation using IDT for Illumina Nextera UD Indexes. Further enrichment and cleanup was performed as per protocols provided by the manufacturer (Illumina Inc). Pooled samples were quantified using Qubit 3.0 or 4.0 fluorometer (Invitrogen Inc.) using the Qubit dsDNA High Sensitivity assay according to manufacturer’s instructions. The fragment sizes were analyzed using TapeStation 4200 (Invitrogen). The pooled libraries were further normalized to 4nM concentration and 25 μl of each normalized pool containing unique index adapter sets were combined in a new tube. The final library pool was denatured and neutralized with 0.2N sodium hydroxide and 200 mM Tris-HCL (pH7), respectively. 1.5 pM sample library was spiked with 2% PhiX. Libraries were loaded onto a 300-cycle NextSeq 500/550 HighOutput Kit v2 and run on the Illumina NextSeq 550 instrument (Illumina, San Diego, CA, USA).

### Midnight Protocol

For Oxford Nanopore sequencing, the Midnight primer kit was used as described by Freed and Silander^55^. cDNA synthesis was performed on the extracted RNA using LunaScript RT mastermix (New England BioLabs) followed by gene-specific multiplex PCR using the Midnight Primer pools which produce 1200bp amplicons which overlap to cover the 30-kb SARS-CoV-2 genome. Amplicons from each pool were pooled and used neat for barcoding with the Oxford Nanopore Rapid Barcoding kit as per the manufacturer’s protocol. Barcoded samples were pooled and bead-purified. After the bead clean-up, the library was loaded on a prepared R9.4.1 flow-cell. A GridION X5 or MinION sequencing run was initiated using MinKNOW software with the base-call setting switched off.

### Genome assembly

We assembled paired-end and nanopore .fastq reads using Genome Detective 1.132 (https://www.genomedetective.com) which was updated for the accurate assembly and variant calling of tiled primer amplicon Illumina or Oxford Nanopore reads, and the Coronavirus Typing Tool^56^. For Illumina assembly, GATK HaploTypeCaller --min-pruning 0 argument was added to increase mutation calling sensitivity near sequencing gaps. For Nanopore, low coverage regions with poor alignment quality (<85% variant homogeneity) near sequencing/amplicon ends were masked to be robust against primer drop-out experienced in the Spike gene, and the sensitivity for detecting short inserts using a region-local global alignment of reads, was increased. In addition, we also used the wf_artic (ARTIC SARS-CoV-2) pipeline as built using the nextflow workflow framework^57^. In some instances, mutations were confirmed visually with .bam files using Geneious software V2020.1.2 (Biomatters). The reference genome used throughout the assembly process was NC_045512.2 (numbering equivalent to MN908947.3).

Raw reads from the Illumina COVIDSeq protocol were assembled using the Exatype NGS SARS-CoV-2 pipeline v1.6.1, (https://sars-cov-2.exatype.com/). This pipeline performs quality control on reads and then maps the reads to a reference using Examap. The reference genome used throughout the assembly process was NC_045512.2 (Accession number: MN908947.3).

Several of the initial Ion Torrent genomes contained a number of frameshifts, which caused unknown variant calls. Manual inspection revealed that these were likely to be sequencing errors resulting in mis-assembled regions (likely due to the known error profile of Ion Torrent sequencers)^58^. To resolve this, the raw reads from the IonTorrent platform were assembled using the SARSCoV2 RECoVERY (REconstruction of COronaVirus gEnomes & Rapid analYsis) pipeline implemented in the Galaxy instance ARIES (https://aries.iss.it). This pipeline fixed the observed frameshifts, confirming that they were artefacts of mis-assembly; this subsequently resolved the variant calls. The Exatype and RECoVERY pipelines each produce a consensus sequence for each sample. These consensus sequences were manually inspected and polished using Aliview v1.27 (http://ormbunkar.se/aliview/).

All of the sequences were deposited in GISAID (https://www.gisaid.org/)^17,18^, and the GISAID accession identifiers are included as part of **Supplementary Table S3**. Raw reads for our sequences have also been deposited at the NCBI Sequence Read Archive (BioProject accession PRJNA784038).

The number and position of the Omicron mutations has affected a number of primers and caused primer drop-outs across a range of sequencing protocols, especially within the RBD (https://primer-monitor.neb.com/lineages). These primer drop-outs have resulted in a number of genomes missing stretches of the RBD, and can affect estimates of mutation prevalence and the determination of the true set of lineage-defining mutations. Given this, bam files of all initial genomes were inspected with IG Viewer to confirm mutation calls where reference calls were suspected to be from low coverage at primer dropout sites^59^.

### Lineage classification

We used the widespread dynamic lineage classification method from the ‘Phylogenetic Assignment of Named Global Outbreak Lineages’ (PANGOLIN) software suite (https://github.com/hCoV-2019/pangolin)^19^. This is aimed at identifying the most epidemiologically important lineages of SARS-CoV-2 at the time of analysis, enabling researchers to monitor the epidemic in a particular geographic region. For the Omicron variant described in this study, the corresponding PANGO lineage designation is BA.1 (lineages v1.2.106). When first characterized the lineage was designated as B.1.1.529 but the emergence of of three sibling lineages to Omicron resulted in the split into sub-lineages (B.1.1.529.1, B.1.1.529.2 and B.1.1.529.3, aliased as BA.1, BA.2 and BA.3). BA.1 contains all the genomes with the original mutational constellation that was designated as Omicron and, at time of writing, is the dominant sub-lineage.

### Recombination testing

To test for the possibility that the Omicron lineage is a recombinant of other SARS-CoV-2 lineages, we used a global subsample of sequences spanning January 2021 to August 2021. Using the NCBI SARS-CoV-2 Data hub^60, 61^, we constructed a dataset containing 221 sequences by randomly sampling five sequences from each month for each continent. No Oceania samples were available from July or August, and no South American sequences were available from July 2021^62^. These sequences were aligned together with a set of five high quality BA.1 and seven BA.2 sequences (representing the known diversity of these clades on 5 December 2021) using MAFFT^63^ with default settings. Whereas 3SEQ^38^, and RDP5^39^ were used to analyse this dataset, a subsample of the 39 most divergent sequences from the dataset was analysed using the GARD recombination detection method^37^. Default program settings were used throughout for recombination analyses, with the exception of RDP5 analysis, in which sequences were treated as linear and the window sizes for the SiScan and BootScan methods (two of the seven recombination detection methods applied in RDP5) were changed to 2000 nucleotides.

### Selection analyses

We investigated the nature and extent of selective forces acting on BA.1 genes encoding individual protein products (a median of 25 unique BA.1 sequences per protein product encoding genome region). A subset of publicly available sequences (from the Virus Pathogen Database and Analysis Resource (ViPR) (https://www.viprbrc.org/) were included as background sequences to contextualize selection signals detectable within the BA.1 lineage at the levels of complete protein product encoding regions, and individual codons (a median of 106 sequences per protein coding region). Sequences were selected quality checked, aligned and subjected to BUSTED, RELAX, MEME, FADE, FEL, and BGM selection analyses (all implemented in HyPhy v2.5.31^64^) using the automated RASCL pipeline as outlined previously^2, 9, 35^.

### Structure modeling

We modelled the spike protein on the basis of the Protein Data Bank coordinate set 7A94, showing the first step of the spike protein trimer activation with one RBD domain in the up position, bound to the human ACE2 receptor^65^. We used the Pymol program (The PyMOL Molecular Graphics System, version 2.2.0) for visualization.

### Phylogenetic analysis

All sequences on GISAID^17, 18^ designated Omicron (n=686; date of access: 7 December 2021) were analyzed against a globally representative reference set of SARS-CoV-2 genotypes (n=12 609) spanning the entire genetic diversity observed since the start of the pandemic. In short, the reference set included: 1. All genomes from Africa assigned to PANGO lineage B.1.1 or any of its descendents, excluding those belonging to a VOC clade; 2. A representative subsampling of global data from the publicly maintained global build of Nexstrain (https://nextstrain.org/ncov/gisaid/global); 3. The top thirty BLAST hits when querying GISAID BLAST for BA.1 and BA.2 sequences. This sampling scheme ensures that we analyze Omicron against the closest variants of the virus. Omicron and reference sequences were aligned with Nextalign^66^. A maximum-likelihood (ML) tree topology was inferred in FastTree^67^ under the following parameters: a General Time Reversible (GTR) model of nucleotide substitution and a total of 100 bootstrap replicates^68^. The resulting ML-tree topology was transformed into a time-calibrated phylogeny where branches along the tree are scaled in calendar time using TreeTime^69^. The resulting tree was then visualized and annotated in ggtree in R^70^.

### Time-calibrated BEAST analysis

To estimate a time-scale and growth rate from the genome sequence data, BEAST v1.10.4^71, 72^ was used to sample phylogenetic trees under an exponential growth coalescent model using a strict molecular clock. All BA.1 assigned genomes from South Africa and Botswana (as of 11 December 2021) were included with some lower coverage genomes removed leaving a total of 553 genomes. The single South African BA.2 (CERI-KRISP-K032307, EPI_ISL_6795834) was included to help stabilize the root of the BA.1 clade but the exponential growth coalescent model was only applied to BA.1 (a constant population size coalescent was used for the rest of the tree). The rate of molecular evolution was estimated from the data. Two runs of 100 million iterations were compared to assess convergence and then post-burnin samples pooled to summarize parameter estimates.

### Birth-death phylogenetic analysis

We analysed the full South Africa & Botswana dataset (n = 552) and the reduced dataset containing only Gauteng Province genomes (n = 277) using the serially sampled birth-death skyline (BDSKY) model^73^, implemented in BEAST2 v2.5.2^74^. To allow for changes in genomic sampling intensity shortly after the discovery of the new lineage, we allowed the sampling proportion to vary with time while keeping all other models parameters constant over the study period. The choice of prior distributions for the model parameters is summarised in Extended Data Table 3.

For each analysis, we used a strict clock model with a fixed clock rate of 7.5×10-4 substitution/site/year and a HKY substitution model. The mean duration of infectiousness was fixed at 10 days^75, 76^. The effective reproductive number, Re, was assumed to be constant with time. The sampling proportion, s, was assumed to be 0 before the collection time of the first sample (2021-11-04) and allowed to change at fixed times that were approximately equidistantly spaced between the first sample and the most recent sample (2021-12-05). The maximum clade credibility (MCC) tree generated from the analysis of the full South Africa and Botswana dataset with a Skygrid coalescent tree prior was used as the starting tree. We kept the subsequent tree topologies fixed such that the resulting MCMC chain only sampled internal node heights.

To assess the robustness of our estimates of Re under different assumptions of temporal variations in the sampling proportion, we repeated the analyses with different numbers of equidistant change-time points (3 or 4). All other model parameters and priors were kept the same.

For each analysis, we ran two independent chains of 1×10e+8 MCMC steps and sampled parameters every 10,000 steps. We used Tracer v1.7^77^ to evaluate MCMC convergence for each of the individual chains (ESS > 200) which were then combined using LogCombiner to obtain the final posterior distribution after removing 10% of each chain as burn-in. The resulting estimates for the doubling time and time of origin are summarised in Extended Data Table 4. The 3-epoch and 4-epoch BDSKY models resulted in similar estimates of the effective reproductive number, Re, for both datasets: 2.74 (95% CI: 2.56-2.92) compared to 2.79 (95% CI: 2.60-2.99) for the South Africa & Botswana dataset, and 4.17 (95% CI: 3.76-4.60) compared to 3.85 (95% CI: 3.50-4.22) for the Gauteng Province dataset.

### Phylogeographic analysis

Markov Chain Monte Carlo (MCMC) analyses were run in duplicate in BEAST v1.10.4^71, 72^ for a total of 100 million iterations sampling every 10,000 steps in the chain. Convergence of runs was assessed in Tracer v1.7.1^77^ based on high effective sample sizes (>200) and good mixing in the chains. Maximum clade credibility trees for each run were summarized in TreeAnnotator after discarding the first 10% of the chain as burn in. Finally, the spatiotemporal dispersal of Omicron was mapped using the R package “seraphim”^78^.

### Estimating transmission advantage

We analyzed 805 SARS-CoV-2 sequences from Gauteng, South Africa, that were uploaded to GISAID with sample collection dates from 1 September - 1 December 2021^17^. We used a multinomial logistic regression model to estimate the growth advantage of Omicron compared to Delta at the time point where the proportion of Omicron reached 50%^79, 80^. We fitted the model using the *multinom* function of the *nnet* package and estimated the growth advantage using the package *emmeans* in R.

The difference in the net growth rates (i.e., the growth advantage) between a variant (Omicron) and the wild-type (Delta) can be expressed as follows^81^:

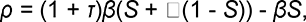

where *τ* is the increase of the intrinsic transmissibility, is the level of immune evasion, *β* is the transmission rate of the wild-type, and *S* is the proportion of the population that is susceptible to the wild-type. This relation can be algebraically solved for *τ* and. We further define *R_w_* = *βSD* as the effective reproduction number of the wild-type with *D* being the generation time. Ω = 1 - *S* corresponds to the proportion of the population with protective immunity against infection and subsequent transmission with the wild-type.

We estimated for different levels of *τ* and *Ω*. To propagate the uncertainty, we constructed 95% credible intervals (CIs) of the estimates from 10,000 parameter samples of *ρ*, *D*, and *R_w_*. 95% credible intervals (CIs) of the estimates from 10,000 parameter samples of We assumed *D* to be normally distributed with a mean of 5.2 days and a standard deviation of 0.8 days^82^. We sampled from publicly available estimates of the daily *R_w_* based on confirmed cases during the early growth phase of Omicron in South Africa (1 October - 31 October 2021; range: 0.78-0.85 (https://github.com/covid-19-Re)^51^.

## Data availability

All SARS-CoV-2 whole genome sequences produced by NGS-SA are deposited in the GISAID sequence database and are publicly available subject to the terms and conditions of the GISAID database. The GISAID accession numbers of sequences used in the phylogenetic analysis, including Omicron and global references, are provided in the **Supplementary Table S1.**

## Code availability

All input files (e.g. alignments or XML files), along with all resulting output files and scripts used in the present study will be made available upon request and publicly shared on GitHub at final publication.

## Acknowledgements

We thank Linda de Gouveia, Amelia Buys, Cardia Fourie, Noluthando Duma, Malusi Ndlovu and other members of the NICD Centre for Respiratory Diseases and Meningitis and Sequencing Core Facility. We thank Nevashan Govender, Genevie Ntshoe, Andronica Moipone Shonhiwa, Darren Muganhiri, Itumeleng Matiea, Eva Mathatha, Fhatuwani Gavhi, Teresa Mashudu Lamola, Matimba Makhubele, Mmaborwa Matjokotja, Simbulele Mdleleni, Masingita Makhubela from the national SARS-CoV-2 NICD surveillance team for NMCSS case data, and Fazil Mckenna, Trevor Graham Bell, Ndivhuwo Munava, Stanford Kwenda, Muzammil Raza Bano and Jimmy Khosa from NICD IT for NMCSS case and test data (in particular, SGTF data). We also thank the following people from the diagnostic laboratories for their assistance: Kubendran Reddy, Lilishia Gounder and Cherise Naicker from NHLS Inkosi Albert Luthuli Central Hospital Laboratory; Stephen Korsman from NHLS Groote Schuur Laboratory; and Annabel Enoch at NHLS Green Point Laboratory. Equally, we thank the global laboratories that generated and made public the SARS-CoV-2 sequences (through GISAID) used as reference dataset in this study (a complete list of individual contributors of sequences is provided in Supplementary Table S3).

The research reported in this publication was supported by the Strategic Health Innovation Partnerships Unit of the South African Medical Research Council, with funds received from the South African Department of Science and Innovation. CA received funding from the European Union’s Horizon 2020 research and innovation programme - project EpiPose (No 101003688). DPM was funded by the Wellcome Trust (222574/Z/21/Z). RC & AR acknowledge support from the Wellcome Trust (Collaborators Award 206298/Z/17/Z - ARTIC network) and AR from the European Research Council (grant agreement number 725422 – ReservoirDOCS). VH was supported by the Biotechnology and Biological Sciences Research Council (BBSRC) (grant number BB/M010996/1). AEZ, JT, MUGK, OGP acknowledge support from the Oxford Martin School. MUGK acknowledges support from the Rockefeller Foundation, Google.org, and the European Horizon 2020 programme MOOD (#874850). MV and the ZARV members, UP was funded through the ANDEMIA G7 Global Health Concept: contributions to improvement of International Health, COVID19 funds through the Robert Koch Institute. The genomic sequencing at UCT/NHLS is funded from the South African Medical Research Council and Department of Science and Innovation; and by the Wellcome Centre for Infectious Diseases Research in Africa (CIDRI-Africa) which is supported by core funding from the Wellcome Trust [203135/Z/16/Z and 222754]. CW and JNB are funded by the EDCTP (RADIATES Consortium; RIA2020EF-3030). Sequencing activities at the NICD were supported by: a conditional grant from the South African National Department of Health as part of the emergency COVID-19 response; a cooperative agreement between the National Institute for Communicable Diseases of the National Health Laboratory Service and the United States Centers for Disease Control and Prevention (grant number 5 U01IP001048-05-00); the African Society of Laboratory Medicine (ASLM) and Africa Centers for Disease Control and Prevention through a sub-award from the Bill and Melinda Gates Foundation grant number INV-018978; the UK Foreign, Commonwealth and Development Office and Wellcome (Grant no 221003/Z/20/Z); the South African Medical Research Council (Reference number SHIPNCD 76756); the UK Department of Health and Social Care, managed by the Fleming Fund and performed under the auspices of the SEQAFRICA project. The genomic sequencing in Botswana was supported by the Foundation for Innovative New Diagnostics and Fogarty International Center (5D43TW009610), NIH (5K24AI131924-04; 5K24AI131928-05), as well in kind support from the Botswana government through the Ministry of Health & Wellness and Presidential COVID-19 Task Force. SM was supported in part by the Bill & Melinda Gates Foundation [036530]. Under the grant conditions of the Foundation, a Creative Commons Attribution 4.0 Generic License has already been assigned to the Author Accepted Manuscript version that might arise from this submission

## Author Contributions

### Genomic data generation

RV, SM, DGA, HT, CS, JG, JE, SG, WTC, DM, BZ, BR, LK, RS, SL, MBM, PS, MM, MM, KM, AM, AI, BM, MSM, JES, NN, GM, SP, TM, UR, YN, CW, SE, TM, WP, LS, UJA, MM, SvW, DT, KD, DH, KM, DD, RJ, AI, DG, PAB, MMN, PNM, JNB;

### Sample collection and metadata curation

RV, SM, DGA, AM, AS, MD, SM, WTC, DM, PS, MC, CJ, LK, OL, KM, NT, NH, NM, KM, AS, AM, MD, ZM, OL, YR, AM, KS, DG, PAB, FT, MV

### Data analysis

HT, CS, RJL, NW, JE, AR, CA, EW, CKW, DPM, VH, RC, JES, MG, SP, AGL, SW, MFB, AEZ, JT, MUGK, OGP

### Study design and data interpretation

RV, SM, DGA, RJL, AR, CA, SG, MM, MM, KM, LK, OL, MSM, KM, CW, OGP, AG, FT, MV, JNB, AvG, TdO

### Manuscript writing

SM, HT, RJL, JG, JE, AR, CA, EW, DPM, JNB, AvG, TdO

All authors reviewed the manuscript

## Additional information

Supplementary Information is available for this paper

Correspondence and requests for materials should be addressed to Professor Tulio de Oliveira, Centre for Epidemic Response and Innovation (CERI), School of Data Science and Computational Thinking, Stellenbosch University, Stellenbosch, South Africa, Reprints and permissions information is available at www.nature.com/reprints

---

**Extended Data Figure 1:**
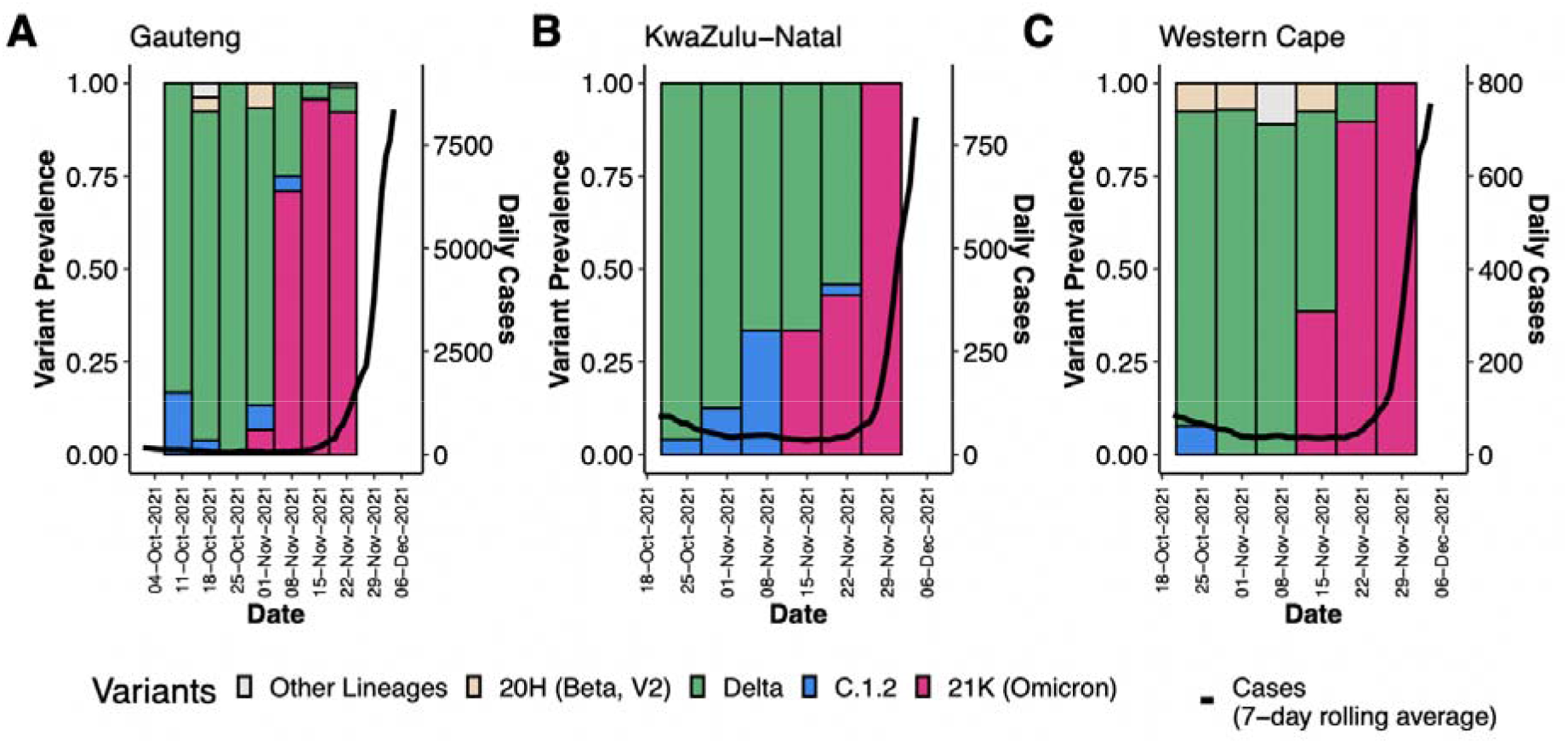
Progression of daily recorded cases and variant proportions in Gauteng (A), KwaZulu-Natal (B) and Western Cape (C) provinces between October and December 2021. A sharp increase in 7-day rolling average of the number of cases is observed in all three of the biggest provinces in South Africa at the emergence of the Omicron variant.

**Extended Data Figure 2:**
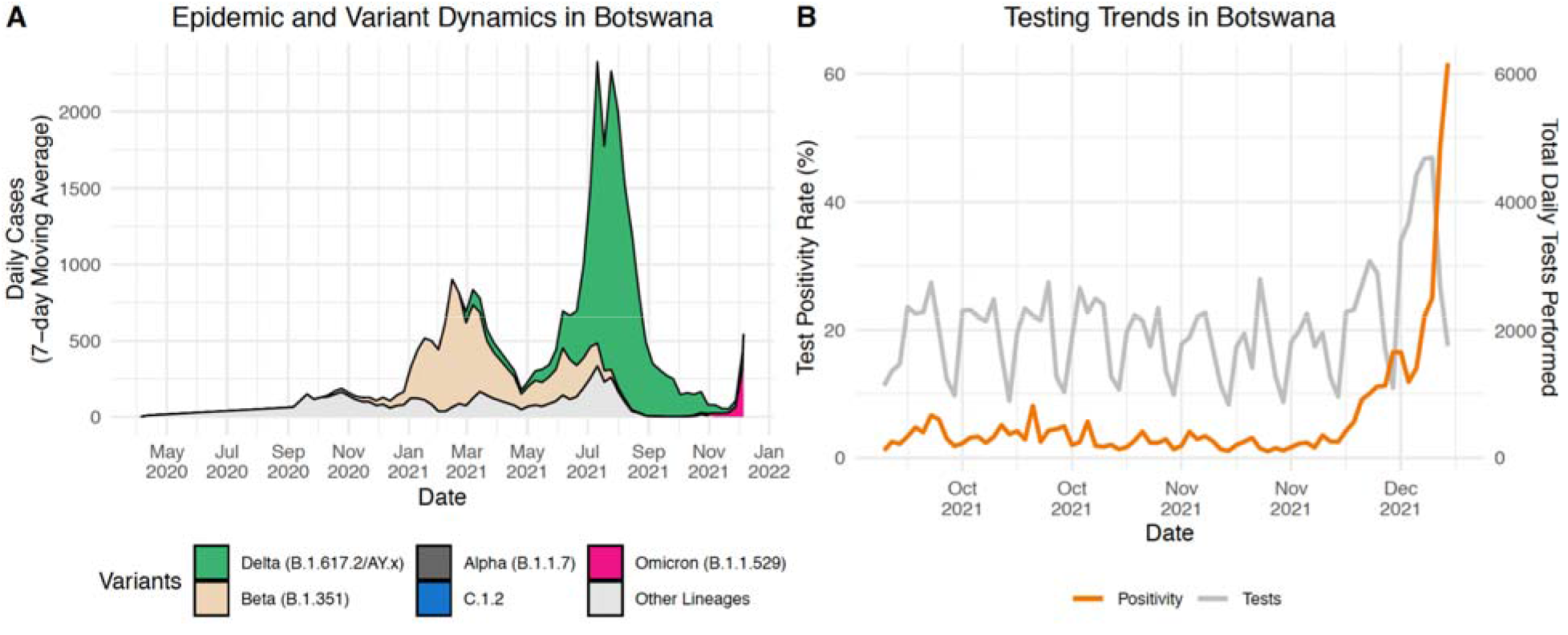
Epidemic Progression in Botswana. A) Epidemic and variant dynamics in Botswana from May 2020 to December 2021, with 7-day rolling average of the number of recorded cases coloured by the proportion of variants as inferred by genomic surveillance data available on GISAID. At the end of November 2021, a big Delta-driven wave was coming to the end and an Omicron wave was starting at the end of November 2021. B) Trends in testing numbers and positivity rates in Botswana between October and December 2021, showing a sharp increase in positivity rate mid-November 2021.

**Extended Data Figure 3:**
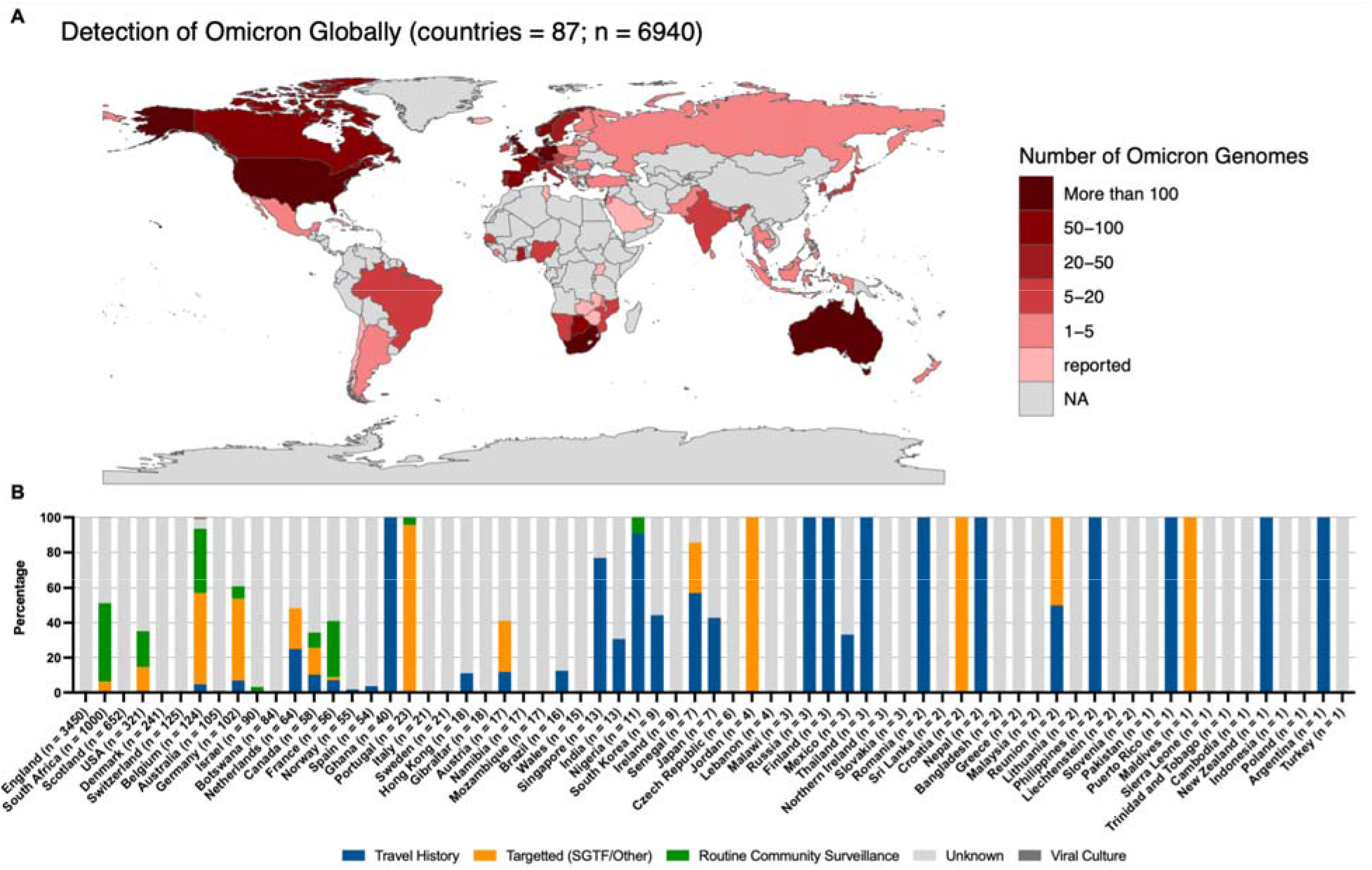
Global distribution of Omicron. (A) Detection of Omicron globally. Shown are the locations for which Omicron genomes have been deposited on GISAID as of December 16, 2021. Those labelled as “reported” referred to the country from which Omicron has been reported to the WHO but there is currently no sequencing data available in GISAID, all data comes from GISAID and the WHO weekly epidemiology report Edition 70 dated December 14, 2021 (https://reliefweb.int/sites/reliefweb.int/files/resources/20211207_Weekly_Epi_Update_69-%281%29.pdf). Countries are coloured according to the number of genomes deposited with the warmer colour representing more genomes. (B) Omicron transmission globally. Shown are countries for which Omicron sequencing data is available on GISAID. Proportion of sequences are coloured according to sample strategy or additional host/location information for either travel history, targeted sequencing (specifically for SGTF, vaccine breakthroughs, outbreaks, contact tracing or other reasons), routine surveillance or unknown if no information has been provided. Countries are ordered by number of sequences available on GISAID as of December 16, 2021.

**Extended Data Figure 4:**
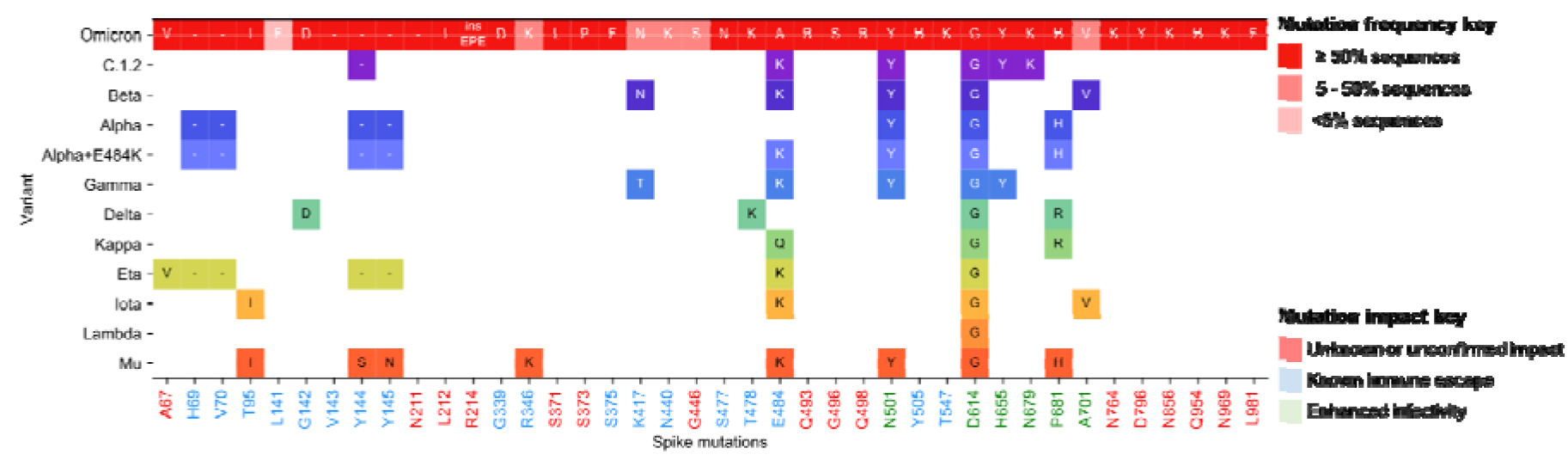
Omicron/BA.1 spike mutations shared with other VOC/VOIs. All spike mutations seen in Omicron/BA.1 are listed at the top in red and coloured according to prevalence. Prevalence was calculated by number of mutation detections / total number of sequences. However, primer drop-outs have affected the RBD region spanning K417N, N440K and G446S, and so it is likely that these mutations may actually be more prevalent than indicated here. For the VOC/VOIs only mutations that are shared with Omicron and seen in ≥50% of the respective VOC/VOI sequences are shown and are coloured according to Nextstrain clade. The mutations listed at the bottom are shaded according to known immune escape (blue), enhanced infectivity (green) or for unknown/unconfirmed impact (red).

**Extended Data Figure 5.**
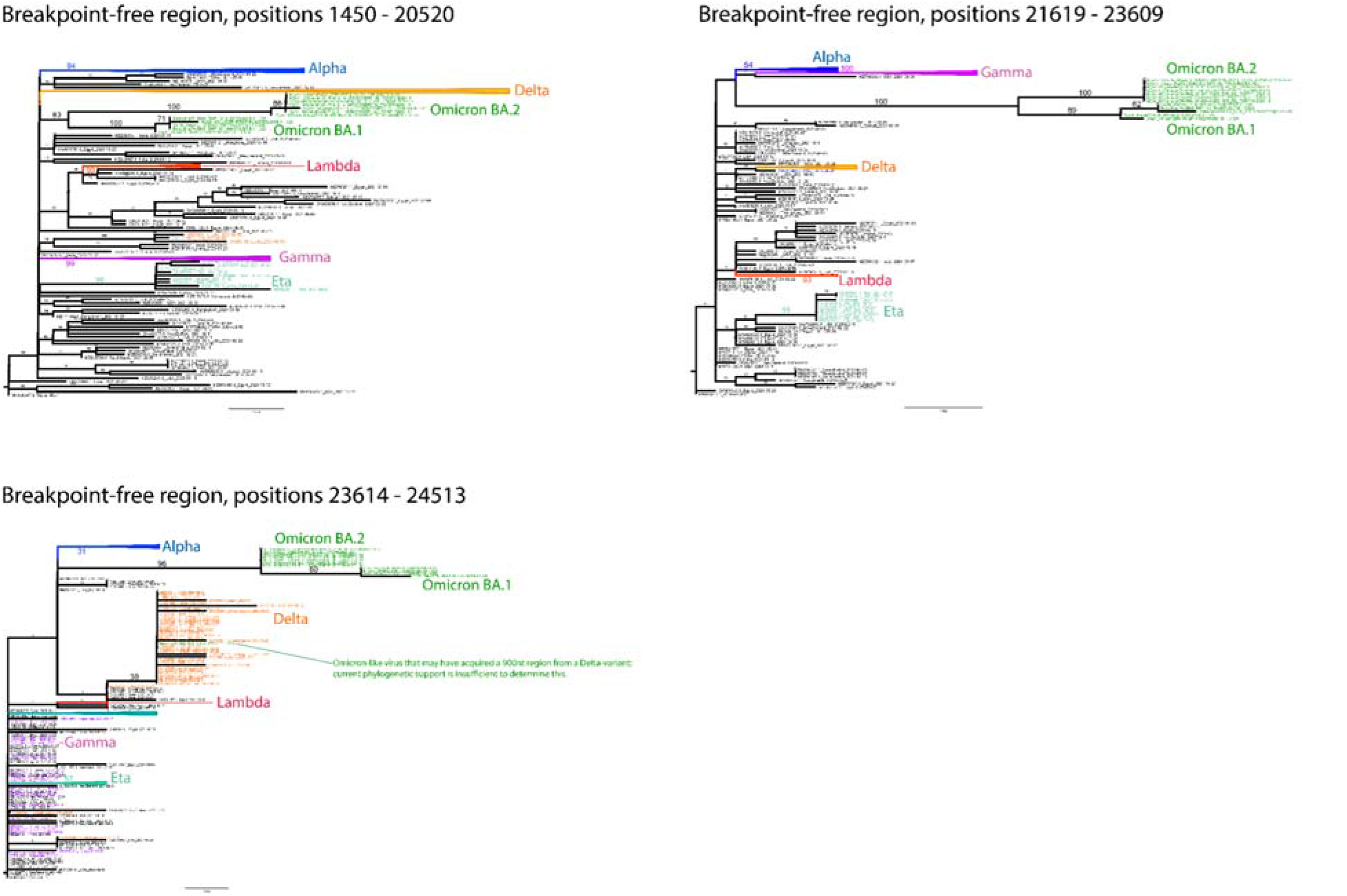
Maximum-likelihood trees (inferred with RAxML v8.2.12 ^83^) for three key breakpoint-free regions (BFR)^84^ of a SARS-CoV-2 genome alignment including global sequences from 2021 (N=221) and 12 sequences of the BA.1/Omicron and BA.2 lineages. All three regions show Omicron as an independently-evolved lineage (bootstrap ≥ 83) with no indication of any regions showing closer/further clustering with non-Omicron lineages. A 900nt segment (in the S2 region) of Botswana/R43B66_BHP_521004487/2021 may have been inserted into this virus through a recombination with a Delta-variant virus, however the clusterin of Botswana/R43B66 with the Delta lineage is supported by a low bootstrap value of 38 due to the short length of this region. Two other candidate BFRs -- one in S2 (positions 26159-27269) and one at the 5’ end of the genome (positions 1-1060) -- showed little genetic diversity and no phylogenetic evidence of recombination. Regions shorter than 900nt had too little genetic diversity to show evidence of phylogenetic clustering. Bootstrap values are shown on branches with relevant values magnified for readability. All trees were rooted on the Wuhan-Hu-1 sequence.

**Extended Data Table 1.** Parameter estimates from BEAST for the full South Africa & Botswana data set and the reduced data set of only Gauteng Province genomes. 95% Highest Posterior Density (HPD) intervals in parentheses.

| Data set | Evolutionary rate<br>$\times 10^{-3}$<br>changes/site/year | BA.1 Time of most recent<br>common ancestor (TMRCA) | Exponential<br>growth rate<br>/day | Doubling time<br>days |
| --- | --- | --- | --- | --- |
| South Africa +<br>Botswana<br>553 Genomes | 1.20 (0.92, 1.49) | 9 Oct 2021 (30 Sep, 20 Oct) | 0.137 (0.100,<br>0.174) | 5.1 (3.8, 7.0) |
| South Africa +<br>Botswana<br>553 Genomes | 0.75 fixed | 1 Oct 2021 (21 Sep, 13 Oct) | 0.139 (0.099,<br>0.183) | 5.0 (4.0, 7.0) |
| Gauteng Province,<br>South Africa only<br>279 genomes | 0.30 (0.08, 0.5) | 16 Sep 2021 (15 Jul, 21 Oct) | 0.222 (0.048,<br>0.396) | 3.1 (1.7, 14.5) |
| Gauteng Province,<br>South Africa only<br>279 genomes | 1.1 fixed | 21 Oct 2021 (16 Oct, 28 Oct) | 0.384 (0.217,<br>0.562) | 1.8 (1.2, 3.2) |

**Extended Data Table 2.** Sites in the Omicron sequences that have been subject to episodic diversifying selection

| Coordinate<br>(SARS-CoV-2) | Gene/ORF | Codon (in<br>gene/ORF) | # of selected<br>branches | AA<br>composition | p-value | Notes |
| --- | --- | --- | --- | --- | --- | --- |
| 1459 | ORF1a | 399 | 1 | K/33, N/1 | 0.0003 |  |
| 2092 | ORF1a | 610 | 1 | S/33, L/1 | 0.0013 |  |
| 2674 | ORF1a | 804 | 1 | P/32, L/1, -/1 | 0.0003 |  |
| 8833 | ORF1a | 2857 | 1 | V/22, -/1, A/1 | 0.0085 |  |
| 10447 | ORF1a | 3395 | 1 | H/15, P/2,<br>Y/1 | 0.0009 |  |
| 10636 | ORF1a | 3458 | 1 | G/15, -/1,<br>F/1, V/1 | 0.0012 |  |
| 17673 | ORF1b | 1402 | 1 | I/29, -/1, V/1 | 0.0060 |  |
| 18264 | ORF1b | 1599 | 1 | R/20, -/1,<br>Q/1, E/1 | 0.0001 |  |
| 18267 | ORF1b | 1600 | 1 | E/20, -/2, T/1 | 0.0005 |  |
| 18270 | ORF1b | 1601 | 1 | E/20, -/2, C/1 | 0.0001 |  |
| 18273 | ORF1b | 1602 | 1 | A/20, -/2, C/1 | 0.0000 |  |
| 21033 | ORF1b | 2522 | 1 | L/10, F/2 | 0.0001 |  |
| 21844 | S | 95 | 2 | I/82, T/3, -/3 | 0.0028 | Clade<br>defining |
| 22576 | S | 339 | 5 | D/77, G/7, -/4 | 0.0000 | Clade |
|  |  |  |  |  |  | defining |
| 22597 | S | 346 | 2 | R/75, K/11, -<br>/2 | 0.0062 | Affects Ab<br>binding |
| 22672 | S | 371 | 2 | L/75, -/7, S/6 | 0.0000 | Clade<br>defining |
| 22684 | S | 375 | 2 | F/75, S/8, -/5 | 0.0061 | Clade<br>defining |
| 22810 | S | 417 | 2 | N/58, -/18,<br>K/12 | 0.0037 | Clade<br>defining |
| 22879 | S | 440 | 3 | K/54, -/27,<br>N/7 | 0.0006 | Clade<br>defining |
| 22897 | S | 446 | 2 | S/56, -/24,<br>G/8 | 0.0038 | Clade<br>defining |
| 22915 | S | 452 | 1 | L/59, -/25,<br>R/4 | 0.0003 | Affects Ab<br>binding |
| 23662 | S | 701 | 2 | A/81, V/6, -/1 | 0.0097 | 501Y<br>Metasignatur<br>e |
| 23851 | S | 764 | 2 | K/75, -/7, N/6 | 0.0083 | Clade<br>defining |
| 23947 | S | 796 | 2 | Y/82, D/5, -/1 | 0.0044 | Clade<br>defining |
| 24127 | S | 856 | 2 | K/82, N/5, -/1 | 0.0006 | Clade<br>defining |
| 24421 | S | 954 | 3 | H/81, Q/7 | 0.0058 | Clade |
|  |  |  |  |  |  | defining |
| 24466 | S | 969 | 2 | K/81, N/6, -/1 | 0.0045 | Clade<br>defining |
| 24502 | S | 981 | 2 | F/79, L/8, -/1 | 0.0048 | Clade<br>defining |
| 28250 | ORF8 | 120 | 2 | L/45, F/35, -<br>/12 | 0.0011 |  |
| 28369 | N | 33 | 0 | -/36, S/6, G/1 | 0.0058 |  |
| 28459 | N | 63 | 2 | D/31, G/6, -/6 | 0.0000 |  |
| 29299 | N | 343 | 1 | D/39, G/3, -/1 | 0.0059 |  |

**Extended Data Table 3.**
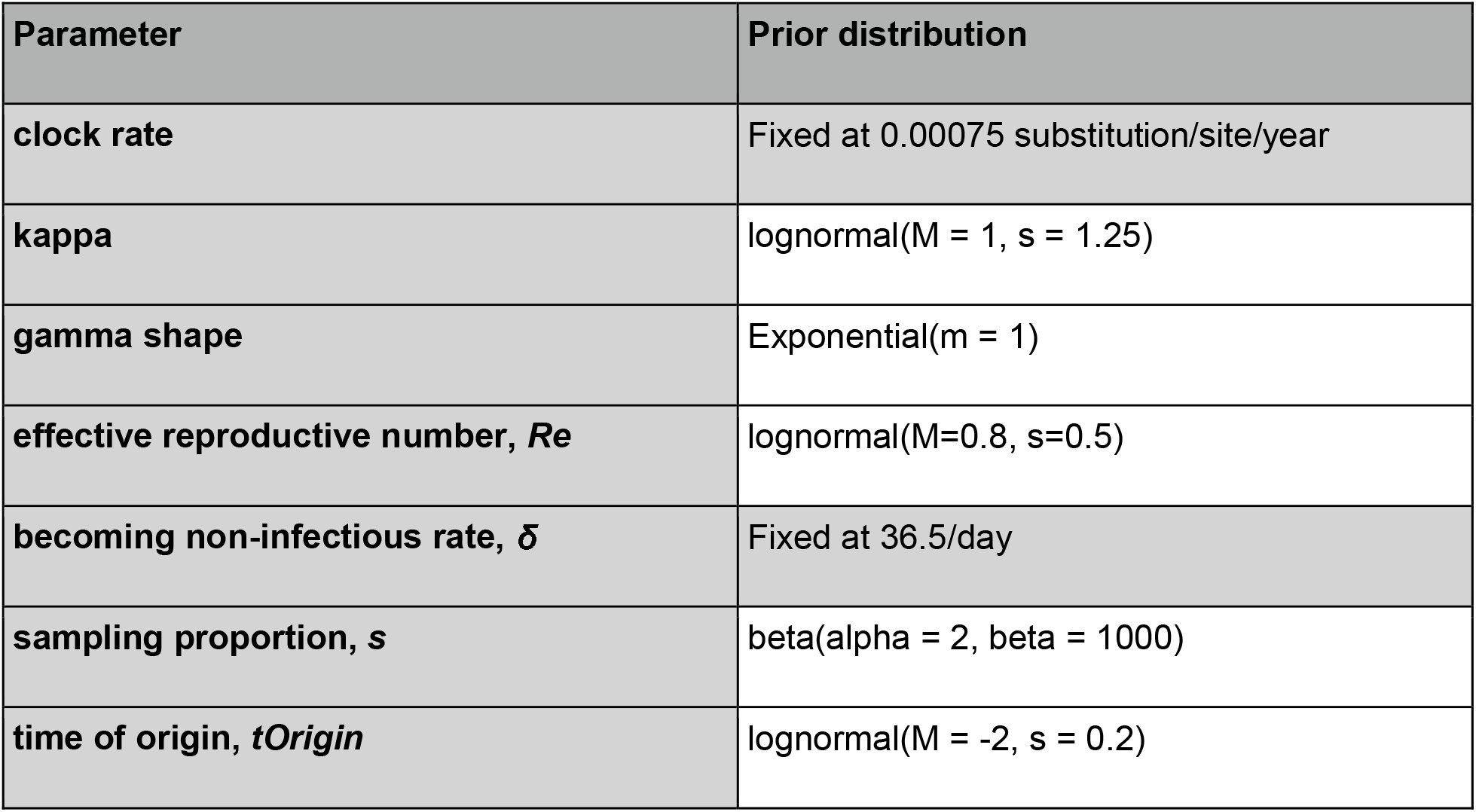
Prior distributions used for the BDSKY analyses. The becoming non-infectious rate was fixed to 36.5/day which corresponds to an infectious period of 10 days.

| Parameter | Prior distribution |
| --- | --- |
| clock rate | Fixed at 0.00075 substitution/site/year |
| kappa | lognormal($M = 1$, $s = 1.25$) |
| gamma shape | Exponential($m = 1$) |
| effective reproductive number, $R_e$ | lognormal($M=0.8$, $s=0.5$) |
| becoming non-infectious rate, $\delta$ | Fixed at 36.5/day |
| sampling proportion, $s$ | beta($\alpha = 2$, $\beta = 1000$) |
| time of origin, $t_{Origin}$ | lognormal($M = -2$, $s = 0.2$) |

**Extended Data Table 4.** Doubling time and time of origin estimates for the full South Africa & Botswana dataset and the reduced dataset of only Gauteng Province genomes under two different assumptions of temporal variation in sampling proportion.

| | doubling time, $\square_d$ | | time of origin, $t_{Origin}$ | |
| --- | --- | --- | --- | --- |
|  | 3-epoch | 4-epoch | 3-epoch | 4-epoch |
| **South Africa + Botswana BA.1 (n=552)** | 3.99<br>[3.61, 4.45] | 3.88<br>[3.47, 4.30] | 2021-10-08<br>[2021-09-27, 2021-10-16] | 2021-10-07<br>[2021-09-26, 2021-10-15] |
| **Gauteng Province only BA.1 (n=279)** | 2.19<br>[1.90, 2.48] | 2.43<br>[2.15, 2.77] | 2021-10-12<br>[2021-10-04, 2021-10-18] | 2021-10-10<br>[2021-10-02, 2021-10-17] |

